# Revealing Individual Neuroanatomical Heterogeneity in Alzheimer’s Disease

**DOI:** 10.1101/2022.06.30.22277053

**Authors:** Serena Verdi, Seyed Mostafa Kia, Keir Yong, Duygu Tosun, Jonathan M. Schott, Andre F. Marquand, James H. Cole, the Alzheimer’s Disease Neuroimaging Initiative

## Abstract

Alzheimer’s disease is clinically heterogeneous, in symptom profiles, progression rates and outcomes. This clinical heterogeneity is linked to underlying neuroanatomical heterogeneity. To explore this, we employed the emerging technique of neuroanatomical normative modelling to index regional patterns of variability in cortical thickness in individual patients from the large multi-site Alzheimer’s Disease Neuroimaging Initiative. We aimed to characterise individual differences and outliers in cortical thickness in patients with Alzheimer’s disease, people with mild cognitive impairment and cognitively normal controls. Furthermore, we assessed the relationships between cortical thickness heterogeneity and cognitive function, amyloid-beta, tau, ApoE genotype. Finally, we examined whether individual neuroanatomical normative maps were predictive of conversion from mild cognitive impairment to diagnosed Alzheimer’s disease. Data on cortical thickness from the 148 brain regions of the Destrieux FreeSurfer atlas was obtained from T1-weighted MRI scans of 1492 participants scanned at 62 different sites. A neuroanatomical normative model was developed to index normal cortical thickness distributions using a separate healthy reference dataset (n= 33,072), employing hierarchical Bayesian regression to predict cortical thickness per region using age and sex. These regional normative models were then fine-tuned to the ADNI dataset after which cortical thickness z-scores per region were calculated, resulting in a z-score ‘map’ for each participant. Regions with z-scores < -1.96 were classified as outliers. Patients with Alzheimer’s disease had a median of 12 outlier regions out of a possible 148. Individual patterns of outlier regions were highly variable, with the highest overlap in the parahippocampal gyrus at only 47% of patients. For 62 regions, over 90% of these patients had cortical thicknesses within the normal range. Patients with Alzheimer’s disease had significantly more outlier regions than people with mild cognitive impairment or controls [*F*(2, 1022) = 95.39), *P* = 2.0 x ×10^−16^]. They were also statistically more dissimilar to each other than were people with mild cognitive impairment or cognitive normal controls [*F*(2, 1024) = 209.42, *P* = 2.2×10^−16^]. Having a greater number of outlier regions was associated with worse cognitive function, CSF protein concentrations and an increased risk of converting from mild cognitive impairment to Alzheimer’s disease within three years (HR =1.028, 95% CI[1.016,1.039], *P* =1.8 ×10^−16^). Individualised normative maps of cortical thickness highlight the heterogeneity of Alzheimer’s effects on the brain. Regional outlier estimates have the potential to be a marker of disease and could be used to track an individual’s disease progression or treatment response in clinical trials.

## Introduction

Alzheimer’s disease is the commonest cause of dementia, being characterised by progressive deterioration in cognitive functioning and independence ^1^. The Alzheimer’s disease spectrum comprises substantial clinical and biological differences between patients recognised in clinical and research criteria. Such differences comprise variations in genetic basis ^2–4^, symptom profile, age-at-onset, trajectory and severity ^5,6^, biomarker readouts (e.g., CSF amyloid-beta levels) ^7^, comorbidities ^8,9^, and in atrophy patterns ^10^. Despite this, conventional statistical analyses in Alzheimer’s disease research focus on group averages. This fundamental statistical assumption posits that Alzheimer’s disease will affect different patients in similar ways ^11^, characterising the ‘average’ patient. To reach the goal of precision medicine for Alzheimer’s we need to look beyond the average and design statistical approaches that reflect the heterogeneity between patients.

Neuroimaging methods are the gold standard of understanding of the *in vivo* brain ^12^. Structural imaging has been described as the ‘imaging workhorse of neurodegeneration’, being commonly recommended in Alzheimer’s disease diagnostic guidelines ^13^. With this in mind, large structural neuroimaging datasets are increasingly available for dementia, such as Alzheimer’s Disease Neuroimaging Initiative (ADNI), Open Access Series of Imaging Studies (OASIS) and National Alzheimer’s Coordinating Center (NACC) ^14–16^ as well as in the general population (e.g., UK Biobank and the Human Connectome Project). These datasets provide the ability to chart variation across cohorts and facilitating individual prediction.

Large neuroimaging datasets have particularly supported the development and application of data-driven methods in Alzheimer’s disease research. This has further revealed that differences in brain structure are very common in patients ^10,17^. Moreover, they have enabled the estimation of disease subtypes from neuroimaging data, as a way to disentangle heterogeneity by grouping patients by distinctive neurobiological and cognitive characteristics ^10,17–20^ and disease progression ^21^. Such subtypes have the potential to stratify patient groups for clinical decision-making, such as regarding treatment strategy, services and therapies tailored to clinico-radiological phenotype and/ or trial enrolment ^22–24^.

Importantly, there are challenges associated with the clinical translation of neuroimaging derived subtypes ^18^. These include validity of subtypes, how distinct subtypes are from each other and how stable subtypes are over the disease course ^17,25^. Moreover, by design, clustering assumes homogeneity within each cluster, despite individual-level variation present, therefore limiting the representation of heterogeneity in the sample ^26^. Individual-level variation is perhaps most apparent in patients with atypical, non-amnestic Alzheimer’s disease who comprise up to a third of young-onset Alzheimer’s disease and face particular barriers to diagnosis and appropriate care ^24^. Arguably, assessing the neurobiology of Alzheimer’s disease at the individual patient level will provide more precise understanding of their disease, likely outcomes and facilitate tailored treatment strategies.

While the concept of patient-centred, individualised precision medicine for dementia is well established, only research has been published using this approach with neuroimaging data in Alzheimer’s. These include techniques based on deep autoencoders ^27^ and machine learning classification ^17^. However, these studies included only limited validation of individualised predictions, did not fully characterise the spatially distributed nature of alterations in Alzheimer’s disease, and failed to assess how neuroanatomical variability related to cognitive performance, CSF biomarkers, disease progression, or genetic factors. Furthermore, these studies did not deal with crucial confounding effects of MRI scanner on heterogeneity.

One technique for capturing individual-level variability in the brain is neuroanatomical normative modelling. This can provide individual statistical inferences with respect to an expected ‘normative’ distribution or trajectory over time. Specifically, neuroanatomical normative modelling adopts this concept by modelling the relationship between neurobiological variables (e.g., neuroimaging features) which are represented regionally and covariates (e.g., demographic variables such as age and sex) to map centiles of variation across a cohort (i.e., z-scores). An individual can then be located within the normative distribution to establish to what extent they deviate from the expected pattern in each measure. By applying this approach to derive neuroanatomical normative models at brain regions, a map can be generated of where and to what extent an individual’s brain differs from the norm ^28,29^. This technique has shown to be suitable for precise mapping of individual patterns of variation in brain structure across multiple psychiatric and neurodevelopmental disorders, thereby parsing the neuroanatomical heterogeneity present ^30,31^. These include attention-deficit hyperactivity disorder ^32^, autism ^33,34^, bipolar disorder and schizophrenia ^35^. Such findings motivate the application of neuroanatomical normative modelling to Alzheimer’s disease ^36^.

Here, we examine individual patterns of variation in brain structure in patients with Alzheimer’s disease using neuroanatomical normative modelling. Using the well-characterised Alzheimer’s Disease Neuroimaging Initiative dataset, we applied a recent implementation of the normative modelling framework, hierarchical Bayesian regression. This technique has shown to efficiently accommodate inter-site variation and provides computational scaling, which is useful when using large studies, or combining smaller studies together, that are acquired across multiple sites in a federated learning framework ^37–39^. We aimed to quantify spatial patterns of neuroanatomical heterogeneity using cortical thickness measures in patients with Alzheimer’s disease, people with mild cognitive impairment and cognitively normal controls, by calculating deviations from normative ranges for each brain region and then identifying statistical outliers. Here we aimed to 1) assess the extent of neuroanatomical variability between individual patients based on overlapping or distinct patterns of outliers in cortical thickness, 2) quantify group differences in between-subject similarity, 3) relate the quantity of neuroanatomical outliers to cognitive performance and disease biomarkers, and 4) examine whether the number of outliers related to subsequent disease progression in people with mild cognitive impairment.

## Materials and methods

### Participants

Participants were derived from two datasets (1) a reference dataset comprised of healthy people across the human lifespan (2) a clinical target dataset which included people with Alzheimer’s disease or mild cognitive impairment in addition to age-matched cognitively normal controls. The reference dataset was made by combining data on healthy people from multiple publicly available sources, including Open Access Series of Imaging Studies (OASIS), Adolescent Brain Cognitive Development (ABCD) study and UK Biobank (UKB) ^16,40,41^, detailed in **Table 2** ^39^. The clinical data used in the preparation of this article was obtained from the Alzheimer’s Disease Neuroimaging Initiative (ADNI) database; http://adni.loni.usc.edu). The ADNI was launched in 2003 as a public-private partnership, led by Principal Investigator Michael W. Weiner, MD. The primary goal of ADNI has been to test whether serial magnetic resonance imaging (MRI), positron emission tomography (PET), other biological markers, and clinical and neuropsychological assessment can be combined to measure the progression of mild cognitive impairment (MCI) and early Alzheimer’s disease (AD). Inclusion criteria were the availability of a baseline T1-weighted MRI, Alzheimer’s disease participants had to meet the National Institute of Neurological and Communicative Disorders and Stroke–Alzheimer’s Disease and Related Disorders Association criteria for probable Alzheimer’s disease ^42^, and were screened to exclude genetic risk for familial Alzheimer’s. MCI participants were reported a subjective memory concern either autonomously or via an informant or clinician, participants had no significant levels of impairment in other cognitive domains. Written informed consent was obtained from all participants before experimental procedures were performed.

### MRI acquisition

For the clinical dataset, T1-weighted images were acquired at multiple sites using 3T MRI scanners. Detailed MRI protocols for T1-weighted sequences by vendor are available online (http://adni.loni.usc.edu/methods/documents/mri-protocols/). The quality of raw scans was evaluated by University of California San Francisco prior to our exclusion criteria. Scans were excluded based on technical problems and significant motion artefacts and clinical abnormalities (https://adni.bitbucket.io/reference/docs/UCSFFSX51/UCSF%20FreeSurfer%20Methods%20and%20QC_OFFICIAL.pdf).

### Estimation of cortical thickness

T1-weighted scans from both the reference and ADNI datasets were processed using a mix of both FreeSurfer versions 5 and 6. Cortical thickness values were generated using the *recon-all* cross-sectional approach ^43^. This cortical thickness algorithm calculates the mean distance between vertices of a corrected, triangulated estimated grey/white matter surface and grey matter/CSF (pial) surface ^44^, which generated the cortical thickness of each region of the Destrieux atlas regions ^45^. This included both mean and median cortical thickness values and 148 regions cortical thickness values for each participant.

Quality control of FreeSurfer processing for the reference dataset relied on automated filtering median-centred absolute Euler number higher than 25, as we have done in prior work ^38,39^. The exclusion of outliers based on Euler numbers has been shown to be a reliable quality control strategy in large neuroimaging cohorts ^46,47^. For ADNI, quality control was based on visual review of each cortical region performed by UCSF. Only scans which passed this quality control were included in the dataset.

### Neuroanatomical Normative modelling

A hierarchical Bayesian regression model was trained on multi-site data to generate normative models per region using the covariates age and sex. This was based on the population variation in the reference dataset (training data), as per Kia and colleagues which adaptively pools parameter estimates across sites via a shared prior over regression parameters across sites ^39^. This simultaneously accounts for inter-site variation and allows sites to borrow strength from one another in a fully Bayesian framework. The advantage of training the models on the large independent dataset, compared to just using ADNI, is that ADNI consists of many sites with small sample sizes. This would result in unstable estimates of normative distributions that could be strongly influenced by outliers or sampling bias. Here, by training on over n=33,000 from only nine datasets (with 60 sites), the model produces much more stable distribution estimates across the entire lifespan. Next, these estimates were conditioned to our specific context, using an adapted transfer learning approach ^39^. The parameters of the reference normative model were recalibrated to the ADNI dataset using 70% of healthy controls per ADNI site, where 70% was used to give stable estimates of the transferred model parameters, given that many of the scan sites in ADNI have quite small sample sizes. The remaining 30% of healthy controls plus MCI and patients with Alzheimer’s disease were used to assess the heterogeneity in neuroanatomical presentation. This process generated regional and mean cortical thickness z-scores for each participant in the clinical dataset, relative to the normative range of the reference dataset. All modelling steps are performed using PCNToolkit (v0.20) (https://github.com/amarquand/PCNtoolkit).

### Statistical analysis

### Group cortical thickness differences

Cortical thickness group comparisons were conducted using t-tests at each region and corrected for multiple comparisons using the False Discovery Rate. Significant p-values were mapped onto the Destrieux atlas using the R package *ggseg* ^48^.

### Outlier definition and statistics

Outliers in terms of low cortical thickness were identified for each region, defined as Z < - 1.96. We only used the lower bound threshold for outliers as we were primarily interested in cortical thinning associated with neurodegeneration. Here the number of outliers were summed across 148 regions for each participant, to give a total outlier count across regions. Linear regression was used to test for group differences in mean cortical thickness Z-score and total outlier count. Additionally, group comparisons at *each* region were conducted using ANOVA and corrected for multiple comparisons using the False Discovery Rate. Hamming distance, a quantitative measure of similarity between binary thresholded cortical thickness outlier vectors was used to measure dissimilarity between individuals. Mean Hamming distances were then compared between groups.

Lastly, to explore spatial patterns of cortical thickness outliers per group, the proportion of participants within each group whose cortical thickness was an outlier (i.e., Z <-1.96) was calculated for each region. This enabled visualisation of the extent to which patterns of outlier regions overlap or are distinct. This was mapped using the Destrieux atlas via the R package *ggseg*. All statistical analyses were implemented in R version 3.6.2.

### Outlier associations with cognitive function and CSF markers

Linear regression adjusting for age and sex were used to examine the relationship between total outlier score and cognitive composite scores (memory or executive function) ^49^ or CSF markers (amyloid-beta and phospho-tau). Linear regression adjusting for age and sex were used to assess the effects of total outlier score, diagnostic group, and their interaction on memory and executive function, measured as ADNI MEM and ADNI EF scores, respectively ^49^. Conversely, we assessed the association between the CSF markers of amyloid-beta and phospho-tau and the total outlier score, adjusting for age and sex. We also use total scores from the Mini-Mental State Examination (MMSE) to stratify outlier maps in both MCI and patients with Alzheimer’s disease groups.

### MCI to Alzheimer’s disease conversion analysis

Follow-up diagnosis status data, up to three years from baseline scan, were obtained from 454 people with mild cognitive impairment. In total, 76 people with MCI at baseline had converted to Alzheimer’s disease within three years. We then ran a survival analysis using Cox proportional hazards regression to assess whether total outlier relating to the risk of converting from MCI to Alzheimer’s disease, controlling for age and sex. We use a Kaplan– Meier plot to illustrate how either a low or high outlier score (split via median) can contribute to the risk of converting.

### Data availability

Statistical analysis scripts are available on GitHub (https://github.com/serenaverdi/ADNI_normative-modelling). The neuroanatomical normative model was generated using the PCNtoolkit software package (https://github.com/amarquand/PCNtoolkit). ADNI data used in this study are publicly available and can be requested following ADNI Data Sharing and Publications Committee guidelines: http://adni.loni.usc.edu/data-samples/access-data/

## Results

### Participants

In the reference dataset, a total of n=33,072 T1-weighted MRI scans were collated across 60 sites, which included males n= 16,170, females n=16,902, with a mean age of 40 years and a range of 8-97 years (This sample is described in detail in Kia et al 2021 and summarised in **Supplementary material, Table 1)** ^39^. The clinical ADNI dataset amounted to total 1,492 participants which were acquired across 62 sites **(Table 1)**. Here 70% of controls were removed from the clinical dataset and were used as a calibration dataset to adapt the normative model to the new sites, these were randomly selected and stratified across sites and gender to make sure all sites and genders are present in the adaptation set. This left a total of 1,027 participants in the final clinical dataset.

**Table 1.** Demographics of ADNI sample. Statistical differences were assessed using ANOVA and chi-squared tests. *MMSE had maximum score of 30.

|  | Controls | MCI | Alzheimer's disease | Total | Statistical differences |
| --- | --- | --- | --- | --- | --- |
| **n** | 621 | 664 | 207 | 1492 | - |
| **Sex (M:F)** | 252:369 | 370:294 | 106:101 | 728:764 | $\chi^2 = 30.01, P = 3.04 \times 10^{-7}$ |
| **Mean ± sd Age (years)** | 72.2 ± 6.8 | 71.9 ± 7.7 | 74.0 ± 8.0 | 72.3 ± 7.4 | $F(2,1489) = 6.72, P = 0.001$ |
| **Age range (years)** | 62.8 – 88.7 | 56.1 – 92.5 | 57.0 – 88.5 | 56.1-92.5 | - |
| **Mean ± sd total MMSE score\*** | 29.06 ± 1.18 | 27.82 ± 1.91 | 22.56 ± 3.19 | 26.94 ± 3.11 | $F(2,1400) = 867.97, P < 2.2 \times 10^{-16}$ |
| **Mean ± sd CSF amyloid-beta (pg/mL)** | 246.55 ± 305.02 | 231.98 ± 275.41 | 178.07 ± 182.37 | 223.00 ± 263.87 | $F(2, 765) = 3.39, P = 0.03$ |
| **Mean ± sd CSF p-tau (pg/mL)** | 34.06 ± 17.96 | 40.82 ± 24.67 | 57.84 ± 32.22 | 43.42 ± 26.73 | $F(2, 765) = 37.11, P < 4.12 \times 10^{-16}$ |
| **ApoE ε4 negative (% = proportion in group sample)** | 385 (69%) | 322 (53%) | 58 (31%) | 765 (56%) | $\chi^2 = 85.92, P < 2.2 \times 10^{-16}$ |

### Patients with Alzheimer’s disease have smaller cortical thicknesses than MCI and control groups

To understand mean effects across the cohort, mean cortical thicknesses were compared across participant groups. Age- and sex-adjusted mean cortical thickness significantly differed between groups overall [*F*(2,1487) = 137.9, *P* = 2.0 × 10^−16]^. Pairwise comparisons (Tukey post-hoc) were all significant [*P*<0.001], with mean cortical thickness being lowest in Alzheimer’s disease (mean = 2.28 SD = 0.13) and highest in controls (mean = 2.42 SD = 0.11), with MCI being intermediate (mean = 2.38, SD = 0.12) **(Supplementary material, Fig.1)**.

Region-level pairwise group comparisons (total of 148 regions) provided evidence cortical thickness measures were on average lower in 133 regions in Alzheimer’s versus cognitively normal controls, in 111 regions in Alzheimer’s versus MCI and in 78 regions in MCI versus cognitively normal controls, after false discovery rate correction **(Supplementary material, Fig.1)**

Next, cortical thickness z-scores, derived from comparison to the normative model, were then compared across participant groups. In this way, we could compare the degree to which each group differed from the separate reference cohort, used to define the normative model. Consistent with comparisons of mean cortical thickness, age- and sex-adjusted z-scores differed between groups overall [*F*(2, 1022) = 69.49), *P* = 2.0 × 10^−16^]. Pairwise comparisons (Tukey post-hoc) were all significant [*P*<=0.003], with Z-scores being lowest in Alzheimer’s disease (mean = -1.27, SD = 1.41), highest in controls (mean = 0.07, SD = 1.04), and intermediate in MCI (mean =-0.28, SD =1.17) (**Supplementary material, Fig.2 A**).

Furthermore, age- and sex-adjusted total outlier counts differed between groups overall [*F*(2, 1022) = 95.39), *P* = 2.0 x ×10^−16^]. Pairwise comparisons (Tukey post-hoc) were all significant [*P*<=0.003], with total outlier counts being highest in Alzheimer’s disease (median = 12, IQR = 28), lowest in controls (median = 2, IQR = 6) and intermediate in MCI (median = 4, IQR = 9) **(Supplementary material, Fig.2 B)**.

Region-level pairwise group comparisons (total of 148 regions) showed higher numbers of outliers in cortical thickness in 79 regions in Alzheimer’s versus cognitively normal controls, in 63 regions in Alzheimer’s versus MCI and 1 region in MCI versus cognitively normal controls, after false discovery rate correction. Region-level group differences in outlier count were mostly evident within temporoparietal and to a lesser extent frontal and occipital regions **(Fig.1, Panel A)**.

**Fig.1.**
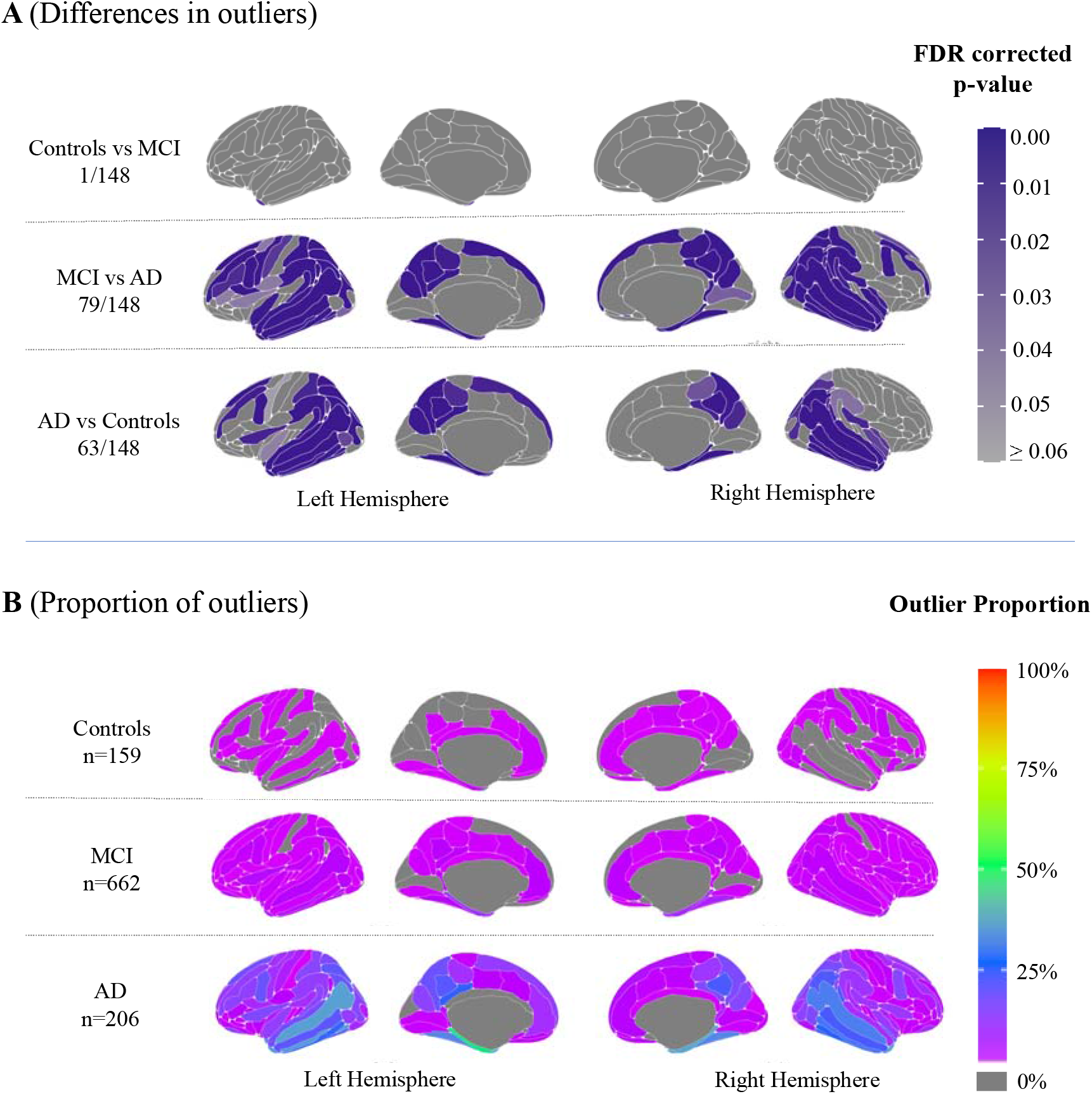
Outlier regions. **(A):** Mapped are significant group differences of region outliers **(B)** Mapped are the percentage of region outliers proportional to participant group.

### Patients with Alzheimer’s disease are less similar to each other than people with MCI or with normal cognition

Hamming distance matrices indicated greater within-group dissimilarity in patients with Alzheimer’s disease, relative to MCI or control participants, who were most similar to each other in spatial patterns of outliers **(Fig.2)**.

**Fig.2:**
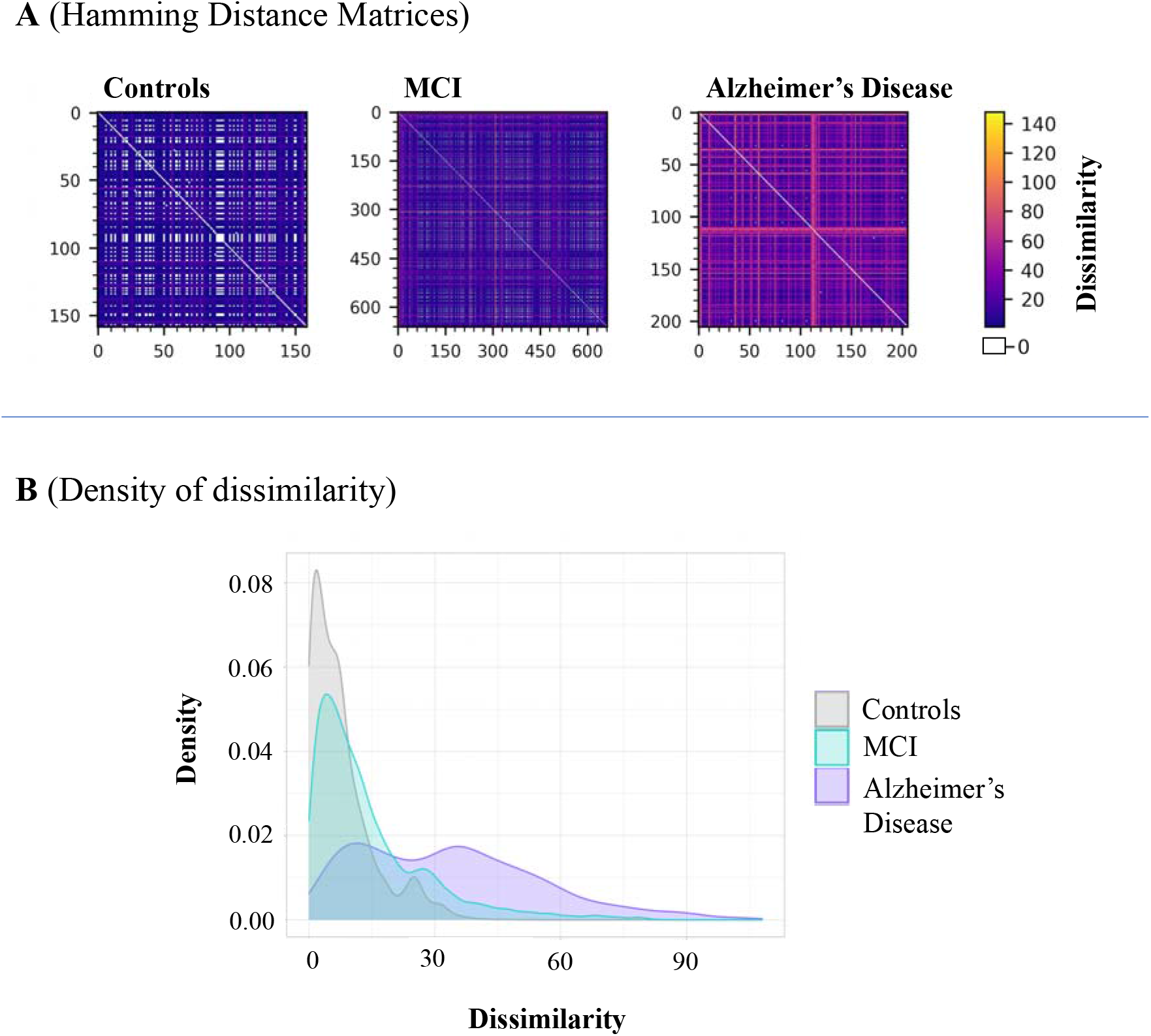
Outlier dissimilarity (A) Outlier distance heatmaps. Both x and y axes represent participants within each group. Yellow indicates higher hamming distance (greater dissimilarity between participants in this brain region), as opposed to if participants are identical in this brain region, the Hamming distance would be 0, represented by white in the colour bar **(B) Outlier distance density:** illustrates the spread of outlier dissimilarity (calculated by Hamming distance) within each group.

Median hamming distance significantly differed between groups overall, [*F*(2, 1024) = 209.42, *P* = 2.2×10^−16^]. Pairwise comparisons (Tukey post-hoc) were all significant [*P*<0.001], with being highest in Alzheimer’s disease (median = 32, IQR = 32), and lowest in controls (median = 6, IQR = 8), with MCI being intermediate (median =10, IQR = 14).

### Patients with Alzheimer’s disease have spatially higher proportions of cortical thickness outliers

The proportion of outliers defined within each group differed in regional patterns between Alzheimer’s, MCI and control groups. This is illustrated in **Fig.1, Panel B** and in **Supplementary material, Fig.3**. For a breakdown of proportions see **Supplementary material, Table 2**. A greater number of regions and a higher proportion of the group were outliers in patients with Alzheimer’s disease, as expected. In fact, 145 regions in the Alzheimer’s group had over the expected 2.5% of patients with an outlier (based on the Z < - 1.96 threshold). The left parahippocampal gyrus was the region with the highest outlier percentage (47% of the Alzheimer’s group). The second highest region outlier proportion was in the right parahippocampal gyrus (36% of the Alzheimer’s group). For the MCI group, 138 regions in the MCI group had outliers (over the expected 2.5% of the group). The left parahippocampal gyrus is the region with the highest outlier percentage (14% of the MCI group). The second highest region outlier proportion was in the right parahippocampal gyrus (12% of the MCI group). For cognitively normal controls, only 66 regions in the control group had outliers above the expected 2.5%. The left occipital temporal lateral sulcus was the region with the highest outlier percentage (6% of controls).

### Outliers are associated with cognitive function and CSF amyloid-beta and phosphorylated tau

Total outlier score across the whole sample was significantly associated with memory performance [*β* = -0.015, *P* = 2.2×10^−16^] and executive function [*β* = -0.02, *P* = 2.2×10^−16^] in a linear regression model. To check for Simpson’s paradox (the association between two variables within a sample), we also model a group by total outlier score interaction term, which was not significant for memory performance [*F*(2, 914) = 1.11, *P* = 0.329] but was for executive function [*F*(2, 914) = 4.04, *P* = 0.017] **(Fig.3, Panel A; B)**. Lower MMSE score showed different spatial patterns of outliers in both MCI (Fig.4, Panel A) and AD (Fig.4, Panel B) groups.

**Fig 3.**
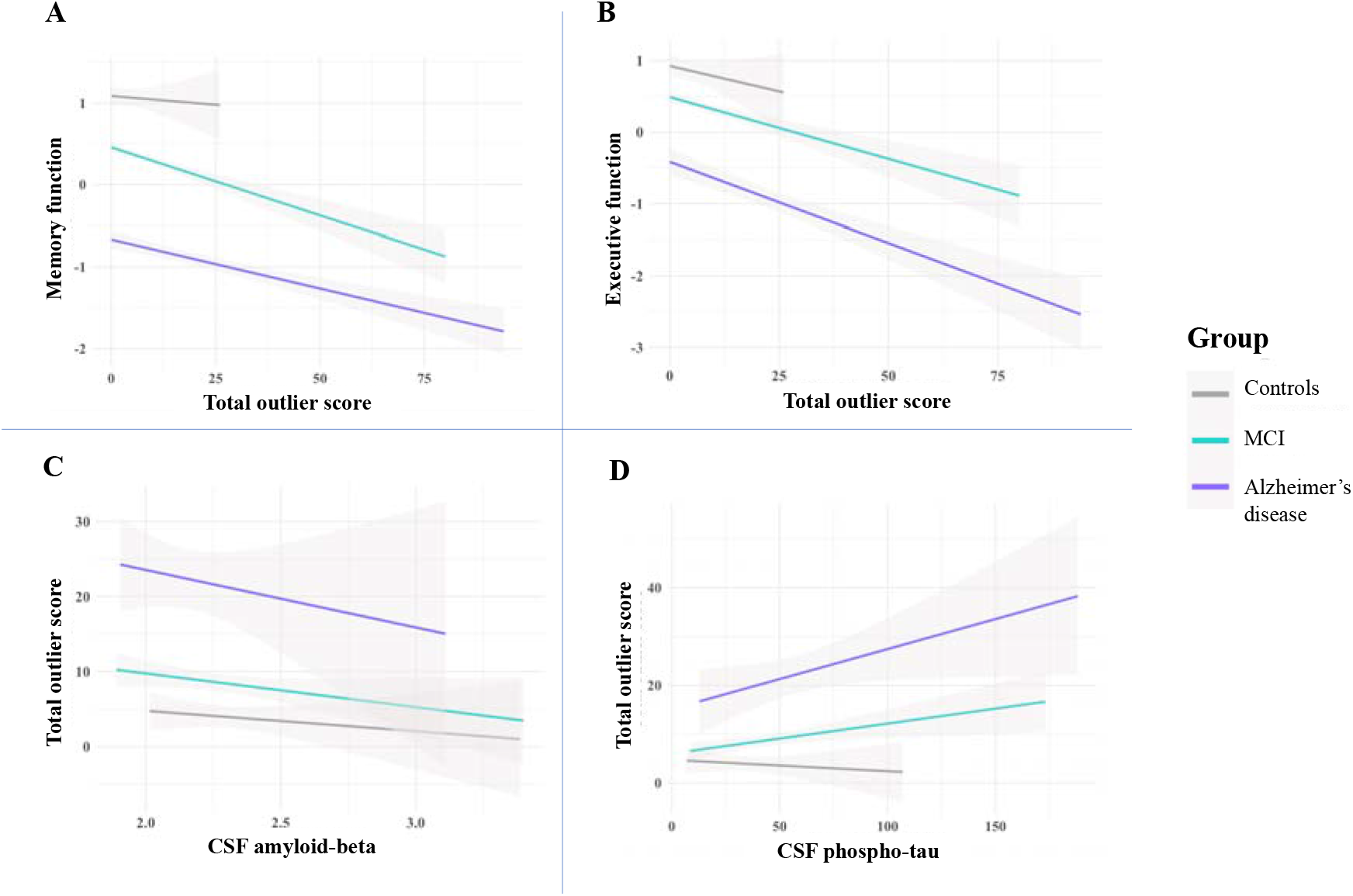
Total outlier count associations with cognitive function and CSF markers. Fitted lines are from a linear regression model per diagnostics group for **(A)** Memory function, **(B)** Executive function, **(C)** CSF amyloid-beta, and **(D)** phospho-tau.

**Fig 4:**
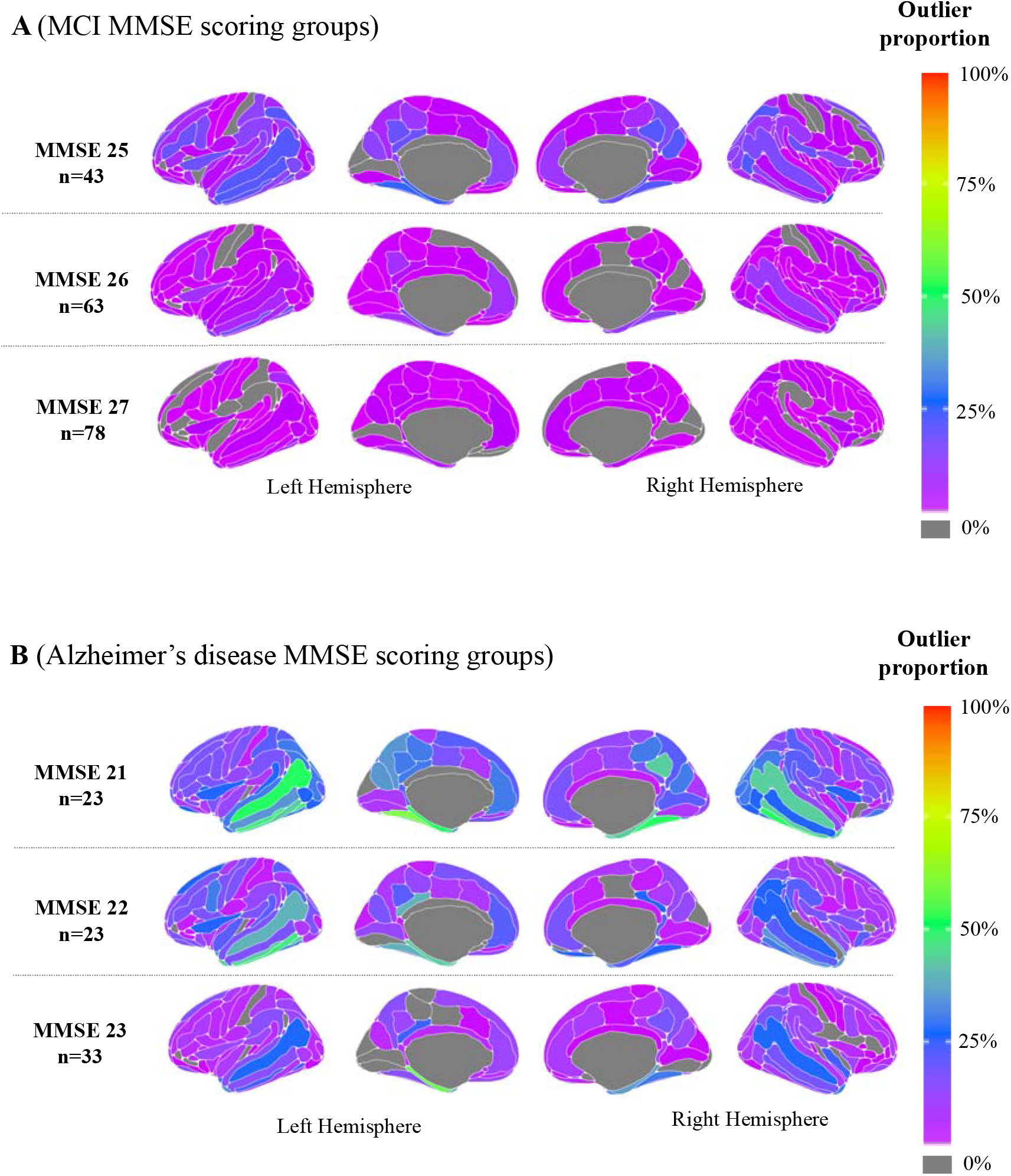
Outlier maps according to MMSE. Mapped are the percentage of region outliers proportional to MMSE scoring subgroup in **(A)** MCI participants **(B)** Alzheimer’s disease participants.

In addition, total outlier score was significantly associated with amyloid-beta [*β* = 0.002, *P* = 0.022] and phospho-tau [*β* = 0.1301, *P* = 1.04×10-8], which was not influenced by either group amyloid-beta [*F*(2, 576) = 0.96, *P* = 0.38] or phospho-tau interaction [*F*(2, 576) = 1.362, *P* = 0.257] **(Fig.3, Panel C; D)**.

### Variability in cortical thickness is not solely due to disease stage or other clinical factors

To explore whether individual differences in outlier maps was driven by disease-related characteristics (such as ApoE genotype, demographics) or by disease stage, we examined sets of participants closely matched for ApoE genotype status, age, sex and MMSE score. **Fig.5** presents four individual female patients with Alzheimer’s all aged 71-72 years, heterozygous for ApoE ε4, with similar MMSE scores, all of whom were amyloid-positive on CSF. These individual patients might be considered similar from biological or clinical perspectives, yet their patterns of outliers in cortical thickness are markedly variable; for example, variously suggesting lateralised (Patient 3) and occipital atrophy (Patient 1). However, MMSE score and age does explain some of the variance in total outlier score [adjusted *R*^*2*^= 0.1942, *P* = 2.2×10^−16^]. Conversely, similar patterns are present within precuneus/post cingulate regions.

**Fig. 5:**
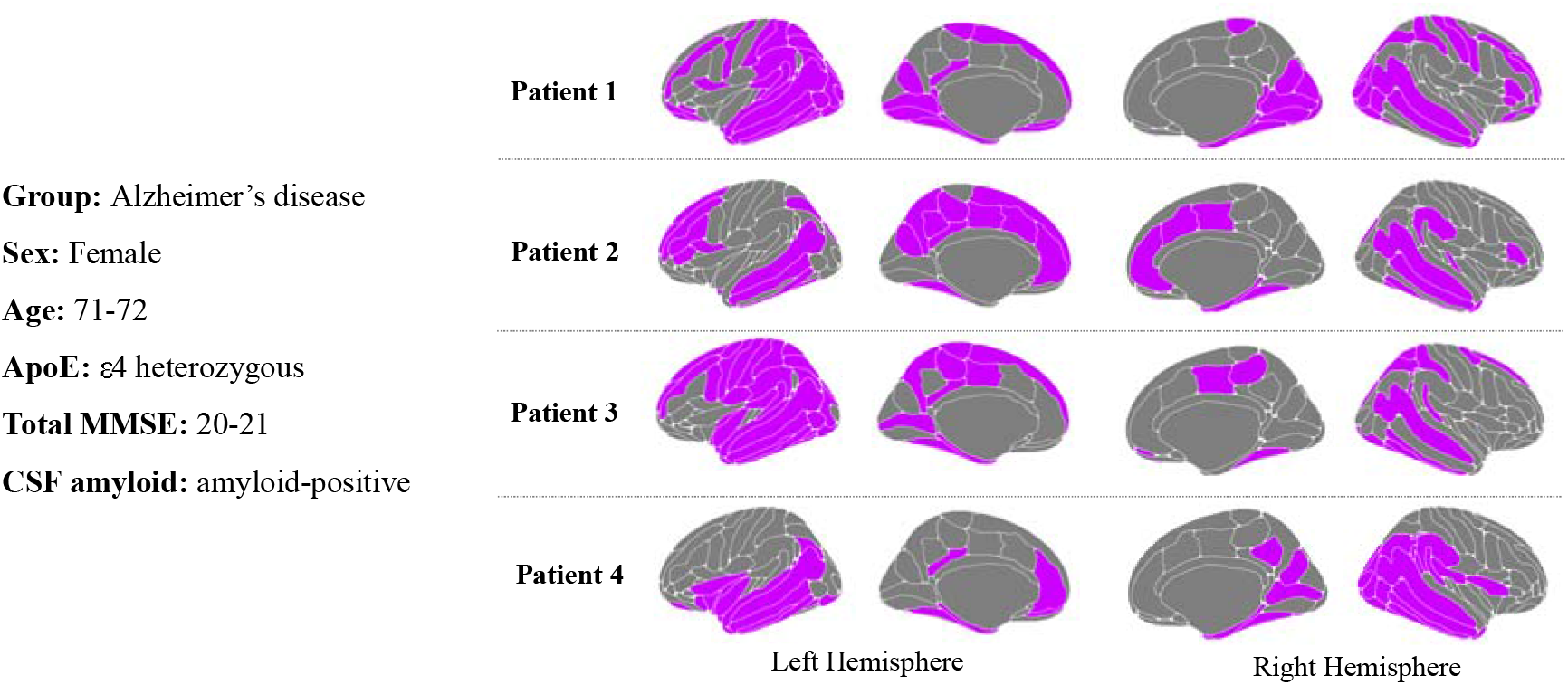
Outlier maps of individuals with similar disease-related characteristics. Outlier regions mapped for individual participants, matched on diagnostic group, sex, age, ApoE ε4, genotype and MMSE score.

**Movie.1:**
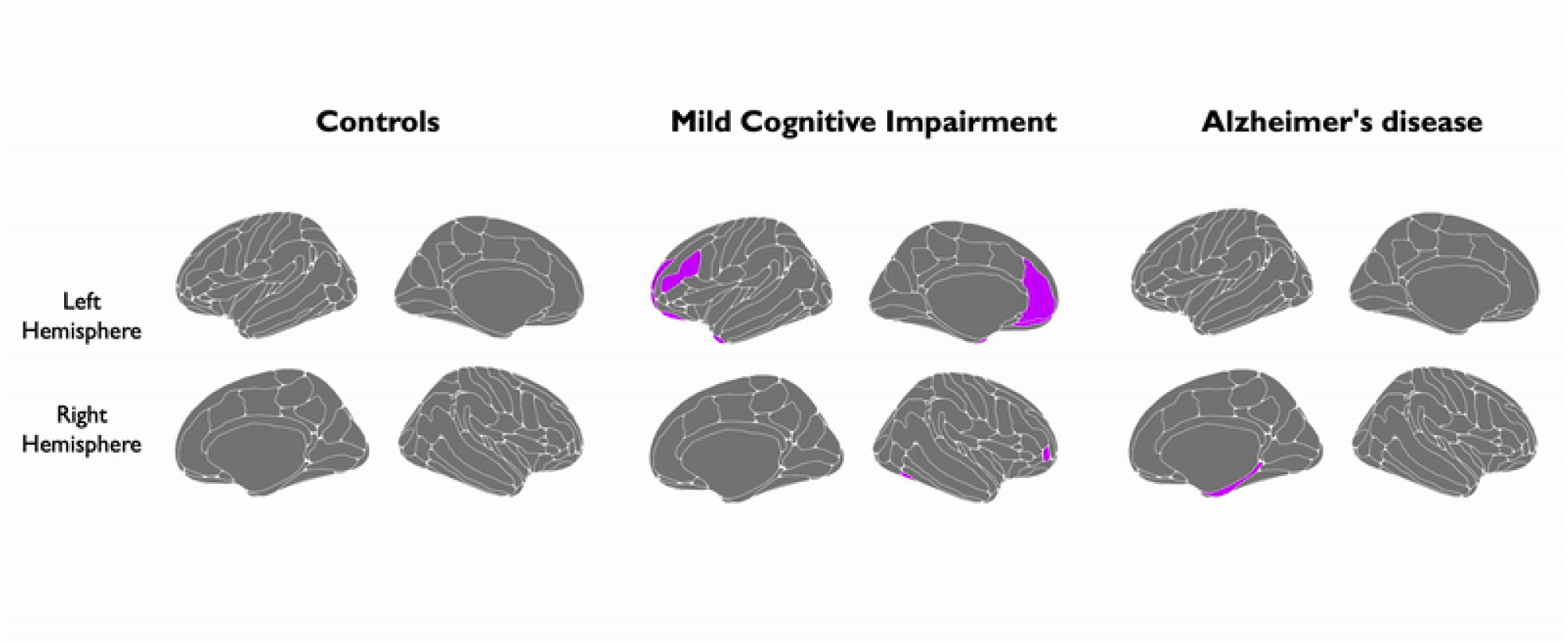
Individual outlier maps (available to play in separate file). Each regional outlier map looping through each participant of this study

### Greater numbers of outliers are associated with risk of conversion from mild cognitive impairment to Alzheimer’s Disease

A survival analysis indicated that for every 10 points of total outlier score, risk of converting from MCI to Alzheimer’s disease within 3 years increased by 31.4% (HR = 1.028, 95% CI [1.016,1.039], *P* = 1.8×10^−16^) **(Fig.6, Panel A)**. This is illustrated within a Kaplan–Meier plot, which shoes how a high outlier score can contribute to the risk of converting in comparison to a low outlier score **(Fig.6, Panel B)**.

**Fig.6:**
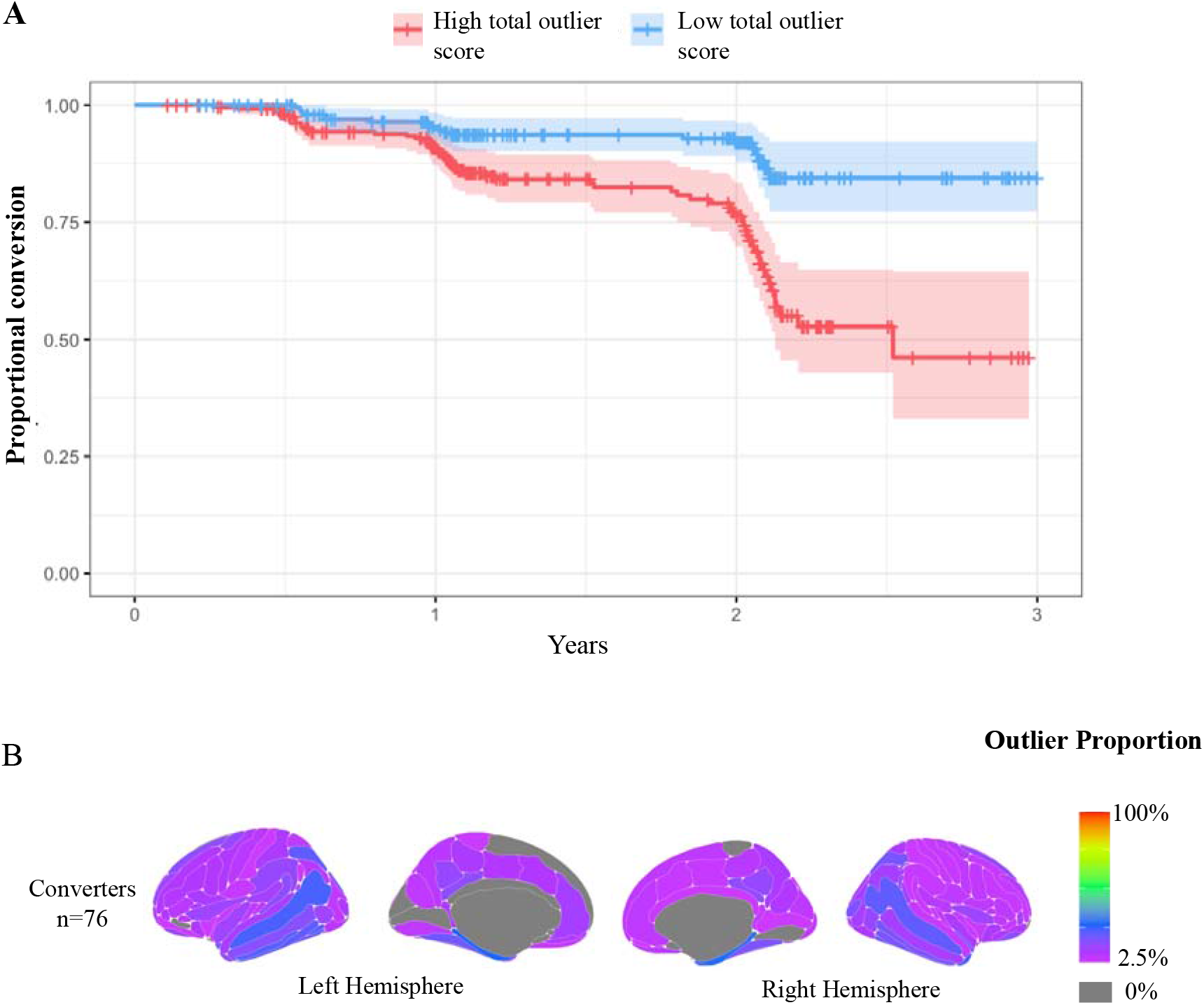
Conversion from mild cognitive impairment to Alzheimer’s Disease. **(A)** Kaplan–Meier plot of MCI to Alzheimer’s disease conversion: the two lines represent a median split of total outlier score, with <4 classed as low outlier score (blue), and ≥4 classed as high outlier score (red). Crosses indicate censoring points (i.e., age at last diagnosis assessment). Filled colour represent the 95% confidence intervals. **(B)** Mapped are the proportion of regional outliers among people with MCI who converted Alzheimer’s disease.

## Discussion

In this study, we defined individual spatial patterns of cortical thickness outliers using a novel neuroanatomical normative modelling approach. Applying this approach to cognitively normal controls, MCI participants and patients with Alzheimer’s disease illustrated that Alzheimer’s disease does not affect different people in a uniform way. Moreover, our analysis provides a way to quantify and visualise these individual differences in patterns of cortical atrophy. The total outlier score (the sum of the total number of outliers an individual has), provides an individualised marker of cortical atrophy for subsequent analyses. Overall, these results provide evidence of, 1) heterogeneous patterns of cortical thickness between patients with Alzheimer’s disease, 2) associations of cortical atrophy with cognitive performance and CSF amyloid-beta and phospho-tau, and 3) the potential of individualised markers of cortical atrophy to predict survival time before conversion from the MCI stage to diagnosed Alzheimer’s disease.

The current findings align with the established understanding of Alzheimer’s disease. We observed a high total outlier count in patients with Alzheimer’s disease, consistent with the evidence of cortical thinning as a consequence of Alzheimer’s disease neuropathology ^50^. Moreover, neuroimaging literature has shown that cortical thinning is significantly associated with poor cognitive performance ^51,52^, a decrease in CSF amyloid-beta and an increase in CSF phospho-tau ^53^, which is also consistent with the present study. Atrophy has also been associated with the risk of progression from MCI to Alzheimer’s disease ^54^, alongside a combination of other biomarkers ^55^. Importantly, all these previous studies examined the correlates of common patterns of cortical atrophy. Conversely, here we were able to specifically consider individual variability in patterns of cortical thickness. This offers precise, personalised neuroanatomical information, as opposed to assessing group average relationships. Potentially, the total outlier score could be used as an individual patient marker of poor brain health. Indeed, related measures have recently been adopted as a clinical measure, i.e., brain volume/thickness patient Z-scores, but has been calculated using different normative modelling techniques ^56,57^. However, these techniques have tended to base their normative population on relatively smaller reference samples; have limited normative modelling pipelines to just whole brain, or within specific regions; did not fully account for site related variation (i.e., site effects) and did not fully relate these to clinical outcomes and cognitive scores

We evaluated group differences in cortical thickness outliers at 148 brain regions, to explore whether consistent neuroanatomical patterns were present across individuals. We observed more outliers in patients with Alzheimer’s disease in temporal regions such as the hippocampus and the cingulate cortex. These are areas known to be sensitive to neurodegeneration in Alzheimer’s disease ^58^, and are responsible for clinical symptoms in Alzheimer’s disease ^59,60^. However, our approach enabled us to go beyond these group-average regional differences. In the Alzheimer’s disease group, the highest proportion of outliers in a single region was less than 50%. This suggests the individual spatial patterns of outliers in Alzheimer’s only partially overlap between patients; if atrophy were homogenous, we might expect 100% of outliers to be here. Heterogeneous atrophy in the temporal lobe is consistent with subtyping studies ^10,61^. Previous efforts have estimated the extent to which multiple distinct atrophy patterns were expressed within each participant, which includes temporal atrophy ^62^. This considerably extends efforts made using autoencoders to calculate individual deviation score in a dataset which includes ADNI patients – here medial temporal regions were also susceptible to scores deviating from the norm ^27^.

Going further, our study reveals that *each* patient not only differs in the number of outliers they have, but the regional patterns of outliers differ markedly. **(Movie. 1)**. The latter is reflected in large levels of dissimilarity between Alzheimer’s disease individuals **(Fig. 2)**. Potentially, one reason for the variable patterns of atrophy is simply disease stage, whereby more atrophy appears with greater disease progression. However, our results indicate that this is not the case, as when closely examining patients of very similar demographics and clinical characteristics, being at a comparable disease stage (e.g., based on MMSE score), heterogenous patterns of cortical atrophy were still present. Crucially, our approach does not preclude the presence of subtypes, although the Z-score maps can be used as input to a cluster algorithm or similar. However, the goal here was to go beyond subtypes, producing informative spatial maps at the individual level.

We also found that lower composite scores of both memory and executive function were significantly associated with higher total outlier score **(Fig. 3)**. While this is consistent with previous studies showing relationships between neuroanatomical variation and cognitive performance,^10,63,64^ this also suggests that individualised measures of neuroanatomy, rather than relying on typical regions (e.g., hippocampus), are sensitive to cognitive function. Potentially, this approach of deriving individualised neuroanatomical measures could benefit clinical trials, where therapeutic effects could be detected across the whole brain, instead of just pre-identified regions of interest.

Cognitively normal controls also showed some outliers suggesting a degree of within-group heterogeneity in unaffected people. The implication of this is that the assumption of homogeneity in case-control studies should be made with caution, even in control groups. Statistical designs for basic research and for clinical trials should better reflect this heterogeneity in brain structure. Although the reference dataset includes over 30,000 individuals, it is still not truly representative of a healthy population ^65^. Also, patients who volunteer for research studies do not necessarily reflect the clinical population; the threshold for contraindications for research scans is higher than clinical scans. Future neuroanatomical normative modelling studies could supplement the reference dataset with MRI scans acquired from routine clinical visits or other less selective sources.

There are some limitations to the current study. The sample taken from this study is cross-sectional, reflecting a snapshot in time, yet heterogeneity has been shown to differ temporally ^66^. Therefore, it will be important for future studies to incorporate serial neuroimaging to define patient-level longitudinal trajectories. In turn, mapping neuroanatomical variability using neuroanatomical normative modelling at different time points has the potential to improve predictions of disease progression or treatment response, at the level of the individual patient. Potentially, data-driven staging methods (e.g., SusStain ^21^) may provide clinically useful information of longitudinal trends of individual heterogeneity whilst taking account of an individual’s disease stage.

The demographic composition ADNI is another issue. In particular, ADNI is comprised of more early-stage dementia participants ^67^. Examining late-stage patients with Alzheimer’s disease may offer interesting insights into the heterogeneity in spatial patterns of atrophy across the disease course. Clinical observations have suggested that late-stage patients with Alzheimer’s disease have widespread atrophy across the brain; therefore, we may hypothesise such patients will have less heterogenous patterns of atrophy. However, regardless of the heterogenous *patterns* of atrophy, the total outlier score can still provide information about the extent of cortical atrophy in a given individual.

Another limitation of ADNI is the limited representation of cognitive domains beyond memory, executive function and language. Between a quarter to a third of the Alzheimer’s disease group exhibit parieto-occipital outliers, comparable to separate parieto-occipital predominant subtypes associated with prominent visuospatial dysfunction ^18^, Further characterisation of how outlier distribution relates to non-memory/executive symptoms may be of particular clinical relevance, for example given the implications of visuospatial dysfunction for diminished autonomy, falls risk and appropriate services ^24,68,69^.

The current approach omitted subcortical and cerebellar regions and could be extended to encompass more of the brain. However, we selected the Destrieux atlas was selected as it contains a higher number of cortical parcellations compared to alternatives in FreeSurfer, to afford better spatial resolution ^45^. Importantly, FreeSurfer is widely used making it easier to amass a large reference dataset of processed data, and readily enabling other research groups to employ the same neuroanatomical normative modelling approach.

## Conclusion

Alzheimer’s disease affects different patients’ brains in different ways. This neuroanatomical heterogeneity aligns with common observations of clinical heterogeneity. While perhaps intuitive, this fact is often overlooked in neuroimaging research and clinical trial design. We provide a quantitative approach to estimate this variability at the individual patient level based on spatial patterns of an individual’s cortical thickness normative deviations. Individualised maps of neuroanatomical outliers were related to cognitive performance and CSF biomarkers. Furthermore, the number of outliers, based on individual patterns, helped predict conversion from MCI to Alzheimer’s disease. These individual neuroanatomical maps, derived from normative models, have the potential to be a marker of Alzheimer’s disease state. These could index disease progression or even to evaluate the effectiveness of potential disease-modifying treatments, tailored to the individual patient.

## Abbreviations

ADNI =: Alzheimer’s Disease Neuroimaging Initiative;
AD =: Alzheimer’s disease;
ApoE =: Apolipoprotein E;
CSF=: Cerebrospinal fluid;
CN =: cognitively normal;
MCI =: Mild Cognitive impairment;
MMSE=: Mini-Mental State Examination

## Acknowledgements

Data collection and sharing for this project was funded by ADNI (National Institutes of Health Grant U01 AG024904) and DOD ADNI (Department of Defence award number W81XWH-12-2-0012). ADNI is funded by the National Institute on Aging, the National Institute of Biomedical Imaging and Bioengineering, and through generous contributions from the following: AbbVie, Alzheimer’s Association; Alzheimer’s Drug Discovery Foundation; Araclon Biotech; BioClinica, Inc.; Biogen; Bristol-Myers Squibb Company; CereSpir, Inc.; Cogstate; Eisai Inc.; Elan Pharmaceuticals, Inc.; Eli Lilly and Company; EuroImmun; F. Hoffmann-La Roche Ltd and its affiliated company Genentech, Inc.; Fujirebio; GE Healthcare; IXICO Ltd.;Janssen Alzheimer Immunotherapy Research & Development, LLC.; Johnson & Johnson Pharmaceutical Research & Development LLC.; Lumosity; Lundbeck; Merck & Co., Inc.;Meso Scale Diagnostics, LLC.; NeuroRx Research; Neurotrack Technologies; Novartis Pharmaceuticals Corporation; Pfizer Inc.; Piramal Imaging; Servier; Takeda Pharmaceutical Company; and Transition Therapeutics. The Canadian Institutes of Health Research is providing funds to support ADNI clinical sites in Canada. Private sector contributions are facilitated by the Foundation for the National Institutes of Health (*www.fnih.org*). The grantee organization is the Northern California Institute for Research and Education, and the study is coordinated by the Alzheimer’s Therapeutic Research Institute at the University of Southern California. ADNI data are disseminated by the Laboratory for Neuro Imaging at the University of Southern California.

## Supplementary material

Supplementary material is available in a separate document.

