## Supplementary Material for "Revealing Individual Neuroanatomical Heterogeneity in Alzheimer’s Disease"

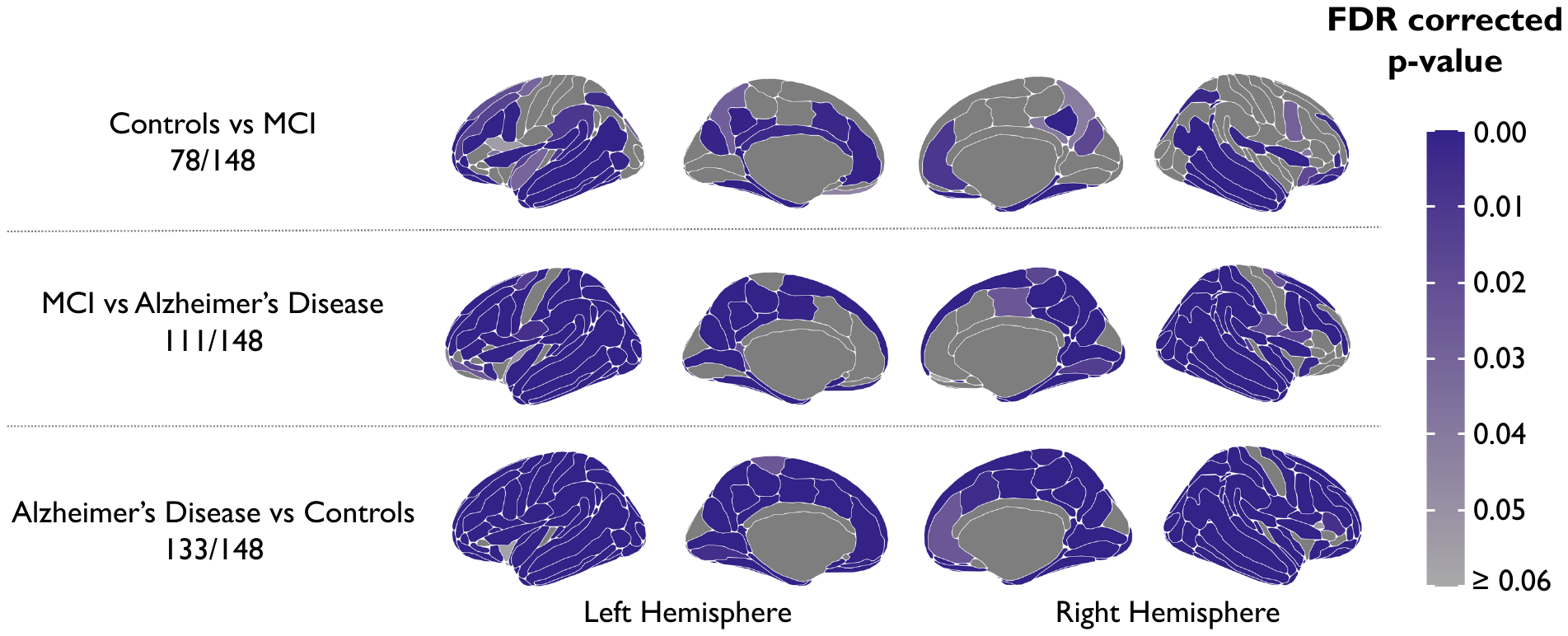

**Supplementary material, Fig.1:** P-value maps to illustrate significant group differences of cortical thickness at each region.

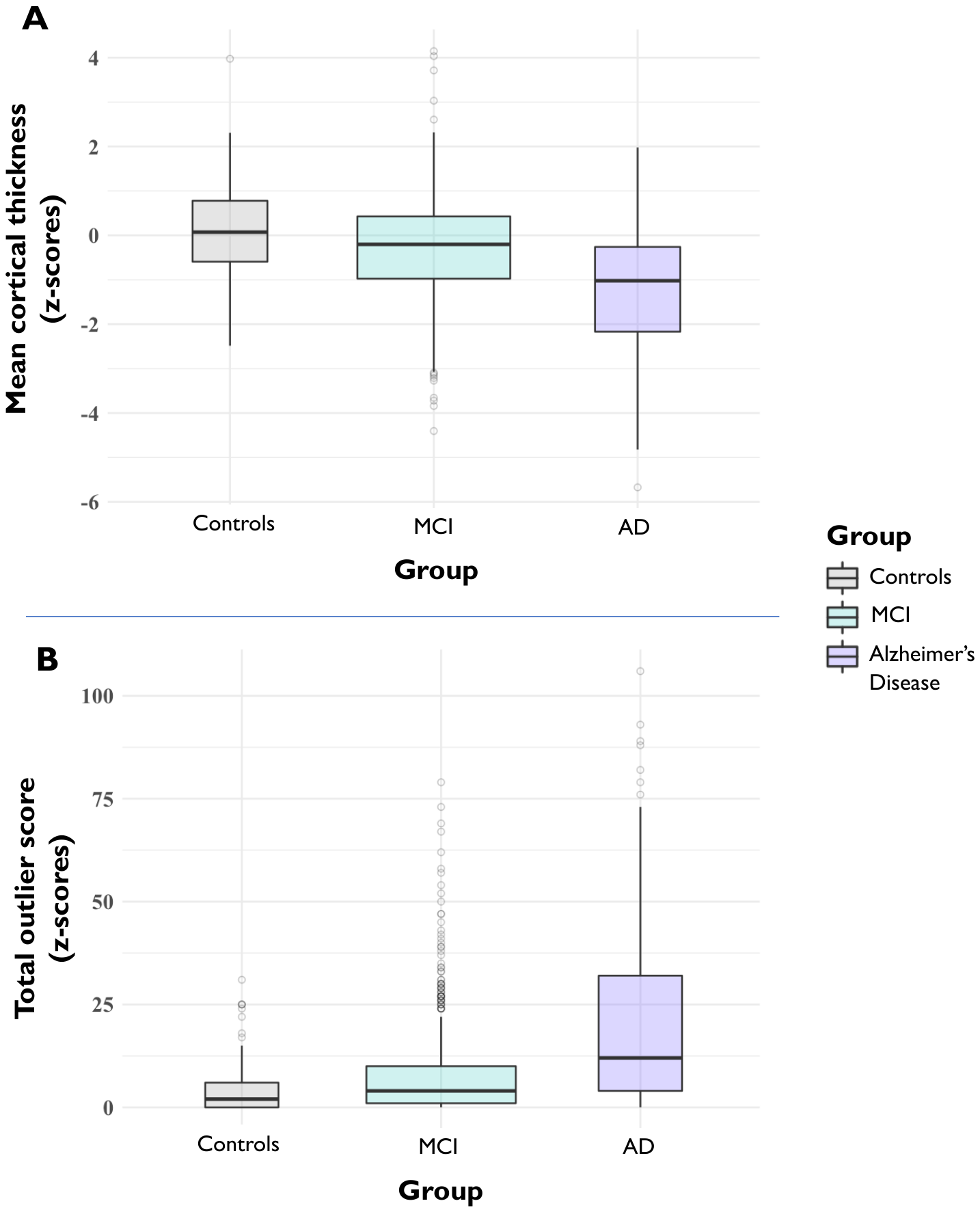

**Supplementary material, Fig.2: (A)** Box plot of group differences of cortical thickness z-scores. Box width is proportional to sample size **(B)** Box plot of group differences of the total outlier count across all regions. **p<0.05; **p<0.001*

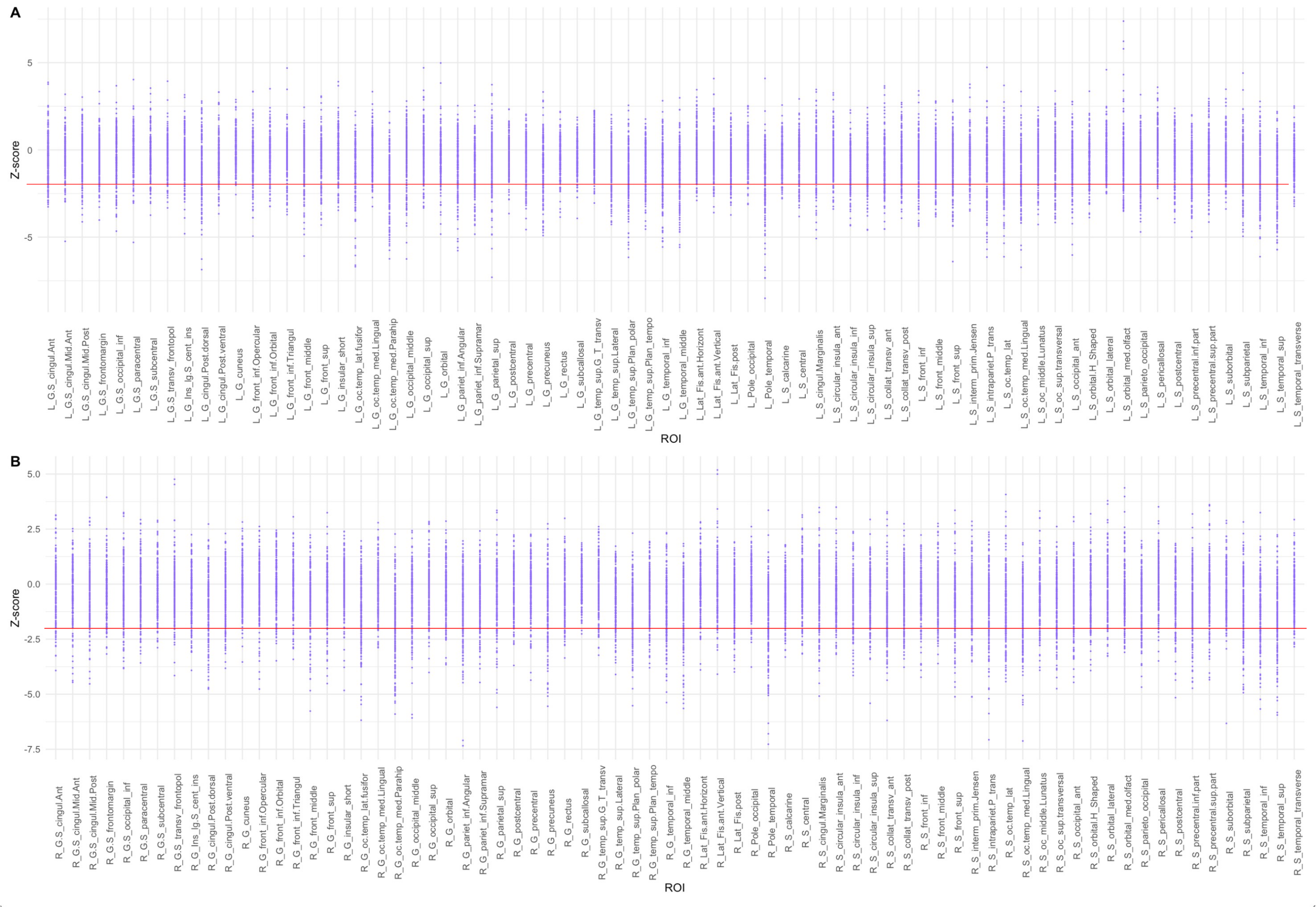

**Supplementary material, Fig.3**: Scatter graph to show distribution of z-scores of Alzheimer’s disease participants. The red line indicates the outlier threshold of -1.96. (A) Left hemisphere (B) Right hemisphere

| **Datasets** | **No. Scans** | **No. Sites** | **Age Range** | **Gender M/F** | **FS Version** |
| --- | --- | --- | --- | --- | --- |
| **ABCD** | 10732 | 29 | 9-11 | 5589/5143 | 6.0 |
| **Cam-CAN** | 647 | 1 | 18-88 | 318/329 | 6.0 |
| **CNP** | 264 | 2 | 21-50 | 152/112 | 6.0 |
| **FCON** | 1021 | 18 | 8-85 | 439/582 | 6.0 |
| **HCP** | 1113 | 1 | 22-37 | 507/606 | 5.3 |
| **OASIS3** | 2044 | 5 | 43-97 | 866/1178 | 5.3 |
| **PNC** | 1514 | 1 | 8-23 | 731/783 | 6.0 |
| **TOP** | 823 | 1 | 17-69 | 435/388 | 6.0 |
| **UKBB** | 14914 | 2 | 44-80 | 7133/7781 | 6.0 |
| **Total** | **33,072** | **60** | **8-97** | **16170/16902** | **-** |

**Supplementary material, Table 1** Demographic breakdown of the datasets which were collated to make the reference dataset. FS = FreeSurfer**,** ABCD = Adolescent Brain Cognitive Development, Cam-CAN = The Cambridge Centre for Ageing and Neuroscience, CNP = The Consortium for Neuropsychiatric Phenomics, FCON = Functional Connectomes Project, HCP = Human Connectome Project, OASIS3 = Open Access Series of Imaging Studies, PNC = The Philadelphia Neurodevelopmental Cohort, TOP (Norwegian datasets), UKBB = UK Biobank

| **Brain region** | **Controls** | **Alzheimer’s Disease** | **MCI** |
| --- | --- | --- | --- |
| L G cingul.Post.dorsal | 2.51572327 | 26.2135922 | 7.55287009 |
| L G cingul.Post.ventral | 3.14465409 | 9.70873786 | 7.85498489 |
| L G cuneus | 0 | 0.97087379 | 1.35951662 |
| L G front inf.Opercular | 1.88679245 | 11.6504854 | 2.87009063 |
| L G front inf.Orbital | 1.25786164 | 6.31067961 | 2.71903323 |
| L G front inf.Triangul | 2.51572327 | 9.22330097 | 3.17220544 |
| L G front middle | 1.88679245 | 15.5339806 | 3.62537764 |
| L G front sup | 1.88679245 | 15.0485437 | 2.41691843 |
| L G Ins lg.S cent ins | 3.14465409 | 10.1941748 | 4.38066465 |
| L G insular short | 1.88679245 | 8.25242718 | 3.32326284 |
| L G oc.temp lat.fusifor | 3.77358491 | 22.815534 | 6.34441088 |
| L G oc.temp med.Lingual | 3.14465409 | 5.82524272 | 3.47432024 |
| L G oc.temp med.Parahip | 5.66037736 | 47.0873786 | 14.1993958 |
| L G occipital middle | 1.88679245 | 17.4757282 | 3.77643505 |
| L G occipital sup | 1.25786164 | 7.76699029 | 1.96374622 |
| L G orbital | 1.25786164 | 7.2815534 | 3.02114804 |
| L G pariet inf.Angular | 0.62893082 | 21.3592233 | 3.92749245 |
| L G pariet inf.Supramar | 0 | 15.5339806 | 4.53172205 |
| L G parietal sup | 1.88679245 | 18.4466019 | 4.07854985 |
| L G postcentral | 1.88679245 | 2.91262136 | 1.35951662 |
| L G precentral | 2.51572327 | 13.1067961 | 3.77643505 |
| L G precuneus | 1.88679245 | 20.8737864 | 4.53172205 |
| L G rectus | 1.88679245 | 6.7961165 | 2.26586103 |
| L G subcallosal | 1.88679245 | 5.82524272 | 1.96374622 |
| L G temp sup.G T transv | 2.51572327 | 2.91262136 | 3.17220544 |
| L G temp sup.Lateral | 0 | 19.9029126 | 4.22960725 |
| L G temp sup.Plan polar | 3.14465409 | 32.038835 | 11.9335347 |
| L G temp sup.Plan tempo | 1.25786164 | 16.9902913 | 5.28700906 |
| L G temporal inf | 0.62893082 | 27.6699029 | 6.64652568 |
| L G temporal middle | 1.88679245 | 22.815534 | 4.83383686 |
| L G.S cingul.Ant | 5.03144654 | 12.6213592 | 8.6102719 |
| L G.S cingul.Mid.Ant | 3.14465409 | 8.25242718 | 4.68277946 |
| L G.S cingul.Mid.Post | 1.25786164 | 8.73786408 | 4.22960725 |
| L G.S frontomargin | 1.88679245 | 9.22330097 | 5.28700906 |
| L G.S occipital inf | 3.77358491 | 11.6504854 | 4.53172205 |
| L G.S paracentral | 1.25786164 | 6.31067961 | 3.17220544 |
| L G.S subcentral | 3.14465409 | 8.73786408 | 2.87009063 |
| L G.S transv frontopol | 1.25786164 | 7.2815534 | 2.71903323 |
| L Lat Fis.ant.Horizont | 1.88679245 | 4.85436893 | 3.62537764 |
| L Lat Fis.ant.Vertical | 3.14465409 | 7.2815534 | 3.62537764 |
| L Lat Fis.post | 3.77358491 | 12.6213592 | 6.79758308 |
| L Pole occipital | 1.25786164 | 2.91262136 | 1.20845921 |
| L Pole temporal | 0.62893082 | 34.4660194 | 11.0271903 |
| L S calcarine | 0 | 5.82524272 | 1.51057402 |
| L S central | 3.14465409 | 10.6796117 | 3.62537764 |
| L S cingul.Marginalis | 1.25786164 | 13.1067961 | 4.53172205 |
| L S circular insula ant | 1.88679245 | 6.7961165 | 3.92749245 |
| L S circular insula inf | 3.77358491 | 15.5339806 | 5.58912387 |
| L S circular insula sup | 3.77358491 | 18.4466019 | 9.36555891 |
| L S collat transv ant | 1.25786164 | 20.3883495 | 8.91238671 |
| L S collat transv post | 1.88679245 | 15.5339806 | 7.09969789 |
| L S front inf | 5.03144654 | 16.0194175 | 6.79758308 |
| L S front middle | 3.77358491 | 13.592233 | 7.55287009 |
| L S front sup | 2.51572327 | 20.8737864 | 5.43806647 |
| L S interm prim.Jensen | 0 | 9.70873786 | 1.05740181 |
| L S intrapariet.P trans | 0.62893082 | 22.3300971 | 8.76132931 |
| L S oc middle.Lunatus | 2.51572327 | 10.6796117 | 6.34441088 |
| L S oc sup.transversal | 2.51572327 | 18.4466019 | 7.25075529 |
| L S oc.temp lat | 6.28930818 | 23.3009709 | 9.06344411 |
| L S oc.temp med.Lingual | 3.77358491 | 32.5242718 | 9.21450151 |
| L S occipital ant | 3.14465409 | 15.5339806 | 7.85498489 |
| L S orbital lateral | 3.77358491 | 4.85436893 | 5.58912387 |
| L S orbital med.olfact | 5.66037736 | 17.4757282 | 5.28700906 |
| L S orbital.H Shaped | 5.03144654 | 16.5048544 | 9.66767372 |
| L S parieto occipital | 1.88679245 | 16.5048544 | 4.83383686 |
| L S pericallosal | 3.77358491 | 2.42718447 | 3.17220544 |
| L S postcentral | 3.14465409 | 16.9902913 | 4.68277946 |
| L S precentral.inf.part | 1.88679245 | 21.3592233 | 7.40181269 |
| L S precentral.sup.part | 3.77358491 | 13.1067961 | 5.58912387 |
| L S suborbital | 2.51572327 | 8.25242718 | 4.53172205 |
| L S subparietal | 3.77358491 | 23.3009709 | 8.4592145 |
| L S temporal inf | 4.40251572 | 34.9514563 | 9.66767372 |
| L S temporal sup | 2.51572327 | 35.9223301 | 9.06344411 |
| L S temporal transverse | 1.25786164 | 9.22330097 | 3.47432024 |
| R G cingul.Post.dorsal | 3.77358491 | 18.4466019 | 7.40181269 |
| R G cingul.Post.ventral | 3.14465409 | 11.1650485 | 3.17220544 |
| R G cuneus | 0.62893082 | 2.91262136 | 2.71903323 |
| R G front inf.Opercular | 3.14465409 | 5.82524272 | 2.87009063 |
| R G front inf.Orbital | 1.25786164 | 6.7961165 | 2.71903323 |
| R G front inf.Triangul | 2.51572327 | 6.7961165 | 3.17220544 |
| R G front middle | 3.14465409 | 12.6213592 | 3.32326284 |
| R G front sup | 1.88679245 | 9.70873786 | 2.41691843 |
| R G Ins lg.S cent ins | 0.62893082 | 11.1650485 | 3.32326284 |
| R G insular short | 1.25786164 | 5.33980583 | 3.62537764 |
| R G oc.temp lat.fusifor | 3.77358491 | 26.2135922 | 8.4592145 |
| R G oc.temp med.Lingual | 1.25786164 | 5.33980583 | 3.77643505 |
| R G oc.temp med.Parahip | 2.51572327 | 36.8932039 | 12.0845921 |
| R G occipital middle | 3.14465409 | 16.5048544 | 4.68277946 |
| R G occipital sup | 2.51572327 | 5.33980583 | 2.71903323 |
| R G orbital | 1.25786164 | 7.76699029 | 3.17220544 |
| R G pariet inf.Angular | 2.51572327 | 24.2718447 | 4.83383686 |
| R G pariet inf.Supramar | 2.51572327 | 13.592233 | 3.77643505 |
| R G parietal sup | 3.14465409 | 14.5631068 | 4.38066465 |
| R G postcentral | 1.25786164 | 5.33980583 | 2.26586103 |
| R G precentral | 4.40251572 | 7.76699029 | 5.43806647 |
| R G precuneus | 3.14465409 | 18.9320388 | 4.98489426 |
| R G rectus | 4.40251572 | 8.25242718 | 3.77643505 |
| R G subcallosal | 1.25786164 | 0.97087379 | 0.45317221 |
| R G temp sup.G T transv | 3.14465409 | 7.76699029 | 4.53172205 |
| R G temp sup.Lateral | 0 | 12.6213592 | 2.71903323 |
| R G temp sup.Plan polar | 4.40251572 | 23.3009709 | 9.51661631 |
| R G temp sup.Plan tempo | 1.88679245 | 11.6504854 | 3.77643505 |
| R G temporal inf | 1.88679245 | 27.184466 | 5.89123867 |
| R G temporal middle | 1.25786164 | 24.7572816 | 4.98489426 |
| R G.S cingul.Ant | 4.40251572 | 9.22330097 | 8.1570997 |
| R G.S cingul.Mid.Ant | 2.51572327 | 7.2815534 | 3.92749245 |
| R G.S cingul.Mid.Post | 5.03144654 | 7.76699029 | 5.13595166 |
| R G.S frontomargin | 2.51572327 | 7.2815534 | 3.62537764 |
| R G.S occipital inf | 5.66037736 | 9.70873786 | 5.74018127 |
| R G.S paracentral | 4.40251572 | 7.76699029 | 2.87009063 |
| R G.S subcentral | 0 | 5.33980583 | 4.07854985 |
| R G.S transv frontopol | 0.62893082 | 6.31067961 | 2.87009063 |
| R Lat Fis.ant.Horizont | 1.25786164 | 4.85436893 | 4.38066465 |
| R Lat Fis.ant.Vertical | 3.14465409 | 5.33980583 | 2.71903323 |
| R Lat Fis.post | 1.25786164 | 13.592233 | 4.53172205 |
| R Pole occipital | 0.62893082 | 4.36893204 | 2.11480363 |
| R Pole temporal | 4.40251572 | 35.9223301 | 11.4803625 |
| R S calcarine | 1.25786164 | 7.2815534 | 1.96374622 |
| R S central | 3.77358491 | 6.7961165 | 3.62537764 |
| R S cingul.Marginalis | 2.51572327 | 14.0776699 | 4.53172205 |
| R S circular insula ant | 4.40251572 | 6.31067961 | 6.49546828 |
| R S circular insula inf | 2.51572327 | 14.5631068 | 4.83383686 |
| R S circular insula sup | 3.77358491 | 14.0776699 | 7.25075529 |
| R S collat transv ant | 1.88679245 | 27.6699029 | 11.4803625 |
| R S collat transv post | 3.14465409 | 16.0194175 | 8.3081571 |
| R S front inf | 1.88679245 | 10.1941748 | 4.38066465 |
| R S front middle | 2.51572327 | 8.25242718 | 6.64652568 |
| R S front sup | 3.77358491 | 13.592233 | 5.58912387 |
| R S interm prim.Jensen | 3.14465409 | 16.5048544 | 5.89123867 |
| R S intrapariet.P trans | 5.03144654 | 19.4174757 | 8.1570997 |
| R S oc middle.Lunatus | 5.03144654 | 12.6213592 | 6.64652568 |
| R S oc sup.transversal | 3.14465409 | 20.3883495 | 6.94864048 |
| R S oc.temp lat | 6.28930818 | 24.2718447 | 11.3293051 |
| R S oc.temp med.Lingual | 5.03144654 | 32.5242718 | 11.3293051 |
| R S occipital ant | 6.28930818 | 18.4466019 | 6.04229607 |
| R S orbital lateral | 3.14465409 | 7.2815534 | 4.98489426 |
| R S orbital med.olfact | 0 | 9.70873786 | 3.77643505 |
| R S orbital.H Shaped | 4.40251572 | 12.1359223 | 7.55287009 |
| R S parieto occipital | 3.77358491 | 17.961165 | 5.13595166 |
| R S pericallosal | 3.14465409 | 3.88349515 | 2.71903323 |
| R S postcentral | 5.03144654 | 14.5631068 | 4.98489426 |
| R S precentral.inf.part | 5.66037736 | 17.4757282 | 3.32326284 |
| R S precentral.sup.part | 5.03144654 | 12.1359223 | 5.13595166 |
| R S suborbital | 1.25786164 | 5.33980583 | 3.32326284 |
| R S subparietal | 4.40251572 | 24.7572816 | 7.85498489 |
| R S temporal inf | 3.77358491 | 29.6116505 | 9.51661631 |
| R S temporal sup | 1.88679245 | 31.0679612 | 7.70392749 |

**Supplementary material, Table 2**: Percentage of outliers defined within each diagnosis group
